# Carotid plaques show more unstable characteristics between 2010-2021 after a prior decade of stabilization

**DOI:** 10.1101/2024.08.26.24312602

**Authors:** Tetiana Motsak, Barend M. Mol, Joost K.R. Hoekstra, Gerard Pasterkamp, Gert J. de Borst, Dominique P.V. de Kleijn

## Abstract

**Background:** Previously we reported a time dependent change in atherosclerotic carotid plaque characteristics, with decreasing destabilising characteristics between 2002 -2011. This observation was considered confirmative with a parallel improved adherence to medication and decrease in overall cardiovascular mortality in Western Europe. In the present study, we investigated if these time dependent changes in plaque characteristics still occurred over the last decade.

**Methods:** Using the Athero Express biobank, atherosclerotic plaques of 1,277 consecutive carotid endarterectomy patients included between 2010 and 2021 were analysed to examine time dependent changes in histological plaque characteristics in intervals of 2 years. These results were compared with our previous time dependent plaque composition data between 2002-2011.

**Results:** In contrast to the period 2002-2011, the period 2010-2021 showed a significant increase in vulnerable plaque characteristics: large lipid cores, intraplaque hemorrhage, macrophages, and calcification. When adjusted for confounders related to these plaque characteristics, such as time to operation and pre-operative type of symptoms, the adjusted odds ratios per 2 years increase in time were 1,177 (95% confidence interval, 1,070-1,293; p<0,001) for calcification, 1,352 (95% confidence interval, 1,229-1,487; p<0,000) for intraplaque hemorrhage, 1,277 (95% confidence interval, 1,159-1,407; p<0,001) for plaques consisting of >40% of fat and 1,388 (95% confidence interval, 1,262-1,528; p<0,001) for macrophages. Use of statins increased in both the 2002-2011 and 2010-2021 period.

**Conclusion:** Our study did not support a further increase in plaque stabilizing features in carotid atherosclerotic plaques between 2010-2021. In contrast, plaques even showed features of destabilisation in the present cohort.

## Introduction

Atherosclerosis is the underlying cause of cardiovascular events like myocardial infarction and stroke with atherosclerotic plaques developing over decades in human life. Atherosclerotic plaque progression can lead to the formation of complex lesions with a large lipid core, thin fibrous cap, high influx of macrophages and intraplaque hemorrhage. These complex lesions can, upon rupture or erosion, result in atherosclerotic cardiovascular disease (ASCVD) related events such as ischemic stroke or myocardial infarction. (1,2) The carotid artery is one of the main atherosclerotic sites where such developments can lead to clinical events. (3–5). While the degree of carotid stenosis is a significant risk factor, the composition of these plaques has independently been associated with accelerated plaque progression, poorer clinical outcomes and can provide additional insights for risk assessment. (1,6–9)

World-wide incidence and mortality of cardiovascular disease is increasing in the last 3 decades (1990-2019) but with large regional differences. (10) In Western Europe, a large decrease in cardiovascular mortality occurs with minor or no changes in incidence. (10) Similar results were found in a large UK study (1980-2013) were prevalence remained unchanged and a declined ASCVD mortality was found together with an increase in hospital admission and ASCVD prescriptions. (11) Much more recently (2006-2019) the SWEDEHEART registry, a large study on myocardial infarction patients, showed that proportion of patients attaining the targets for blood pressure and LDL-C increased significantly. (12) This registry, however, also showed that central obesity, diabetes and patients reporting inadequate levels of physical activity also increased in the last decade. (12)

With the decrease in ASCVD mortality in Western-Europe in the background, time-dependent changes in atherosclerotic plaque characteristics were studied in carotid endarterectomy patients between 2002 -2011 (13). This revealed that carotid atherosclerotic plaques showed a time-dependent change in plaque composition characterized by a decrease in, what are considered, destabilizing plaque characteristics like plaque thrombosis, plaque atheroma, plaque calcification and plaque macrophages. (13) This is in line with decreased mortality and increased ASCVD prescriptions. (10,11) Time-dependent changes in plaque composition in the last decade, however, have not been studied yet despite an increase in diabetes and obesity and less physical activity. (12)

In the present study, we investigated the hypothesis that these time dependent changes in plaque characteristics still occurred over the last decade. Therefore, we examined carotid plaques collected in the Athero-Express biobank (14) to study temporal changes in plaque composition between 2010-2021 with our previous time dependent plaque composition data between 2002-2011.

## Methods

### Study Population

For this study, atherosclerotic plaques from the Athero Express biobank were used. The Athero Express biobank is an ongoing, prospective, longitudinal biobank study and collects atherosclerotic plaques and plasma of both carotid- and femoral endarterectomy patients. An extensive study design has been published earlier (14). Written informed consent was obtained from all included patients and the medical ethics committees of both the University Medical Center Utrecht and St Antonius Hospital Nieuwegein approved the study. For patients included from 2002 to 2011, data was used from our previous Athero Express publication on trends in plaque composition(13). New data was included in this study from consecutive patients included in the Athero Express between 2010 up to 2021 who underwent carotid endarterectomy (CEA). Patients were excluded from the study if informed consent was lacking. Indication for surgery was based on international guidelines for symptomatic and asymptomatic carotid stenosis (15,16). All indications were reviewed by a multidisciplinary team of vascular specialists before surgery. Patient data were obtained via standardized questionnaires and preoperative admission charts. The full Athero Express study is conducted in accordance to the declaration of Helsinki and STROBE reporting guidelines for cross sectional studies was followed. (17,18) One author had full access to all the data in this study and takes responsibility for its integrity and data analysis.

### Plaque Processing and Assessment

Atherosclerotic plaques of patients were harvested during CEA according to a standardized and previously reported protocol.(14,19) Briefly, carotid plaques were divided into segments of 5-mm thickness. The section with the largest plaque burden was classified as the culprit lesion and subjected to immunohistochemical staining. Macrophages, smooth muscle cells (SMC), thrombus, collagen, and calcifications were scored semi-quantitatively at a x40 magnification, according to the following criteria: no (1) or minor (2) staining along part of the luminal border of the plaque or a few scattered spots within the lesion; moderate (3) or heavy (4) staining was scored when along the entire luminal border or evident parts within the lesion staining was present. These categories were grouped into no/minor and moderate/heavy for trend descriptions and logistic regression analyses. Size of the lipid core was, as was previously done, visually assessed using haematoxylin-eosin (HE) stains and picro Sirius red stain with polarized light with a cut off at 40% of the plaque area. Intraplaque hemorrhage was defined as any thrombus staining assessed with HE stains. All of the histologic scorings were performed by the same dedicated investigators during the entire study period.

### Definitions of Risk Factors and Medical Treatment

For baseline characteristics, all patients were asked to complete an extensive questionnaire which was checked with medical records. Height and length were measured before surgery in the outpatient clinic to calculate the BMI. Blood withdrawal was performed at baseline to assess creatinine level, which was used to calculate the estimated glomerular filtration rate with the Modification of Diet in Renal Disease formula (MDRD) (20). Patients were considered smokers if they reported to be smoking until the year of inclusion. Diabetes mellitus was restricted to those patients receiving medical treatment including insulin or oral glucose-lowering drugs. All medication was registered at baseline, including statins, use of antihypertensive drugs including β-blockers, angiotensin-converting enzyme inhibitors, angiotensin II antagonists, and diuretics. Furthermore, prescription of antithrombotic and anticoagulant medication was registered. The index event was registered as transient ischemic attack (TIA), stroke, or ocular symptoms (including amaurosis fugax or retinal infarction) or alternatively as asymptomatic if no neurological symptoms were present within 6 months of the operation. In addition, the time interval between the last symptomatic event and surgery was registered.

### Follow-Up and Outcome

Patients included in the Athero-Express study were asked to fill in a follow-up questionnaire after 1, 2, and 3 years regarding potential cardiovascular events. If the questionnaire was answered positively, further research was performed to investigate the potential outcome of the event. If the patient did not respond, the general practitioner was contacted if patient gave their consent on their informed consent form. The composite end point included any death of presumed vascular origin (stroke, myocardial infarction, sudden death, or other vascular death), ischemic stroke and myocardial infarction.

### Statistical Analyses

To investigate time trends with sufficient sample sizes in different time periods, we studied plaque and patient characteristics over 2-year intervals from 2010 to 2021. To compare baseline and plaque characteristics over time, Pearson’s Chi-squared tests, Fisher’s exact test and Kruskal-Wallis rank sum test were performed when applicable. To compare variables between the entire 2002 – 2011 and 2010 – 2021 periods, Mann-Whitney U Tests and Chi-square tests were used when applicable. To assess changes in time for histological plaque characteristics, a logistic regression analysis was performed. To correct for potential confounders, the multivariate logistic regression analysis was corrected for baseline characteristics with incomparability over time (*p-*value for selection <0.20). For this analysis, plaque characteristics were dichotomized. For macrophages, calcification, SMC’s and collagen into no/minor and moderate/heavy staining. Fat was binned below or above 40% of plaque volume and intraplaque hemorrhage either as present or absent. Two sided p-values of <0.05 were considered statistically significant and all statistical analyses were performed with RStudio version 4.2.1. (R Foundation, Vienna, Austria).

## Results

To investigate if patients undergoing CEA are different within the period 2010-2021 and 2002-2011 we compared baseline patient characteristics including medication and plaque characteristics over both periods. This was summarized in 2 tables (Table 1 baseline characteristics and medication; Table 2 plaque characteristics) that includes both periods (2002-2011 and 2010-2021)

**Table 1.**
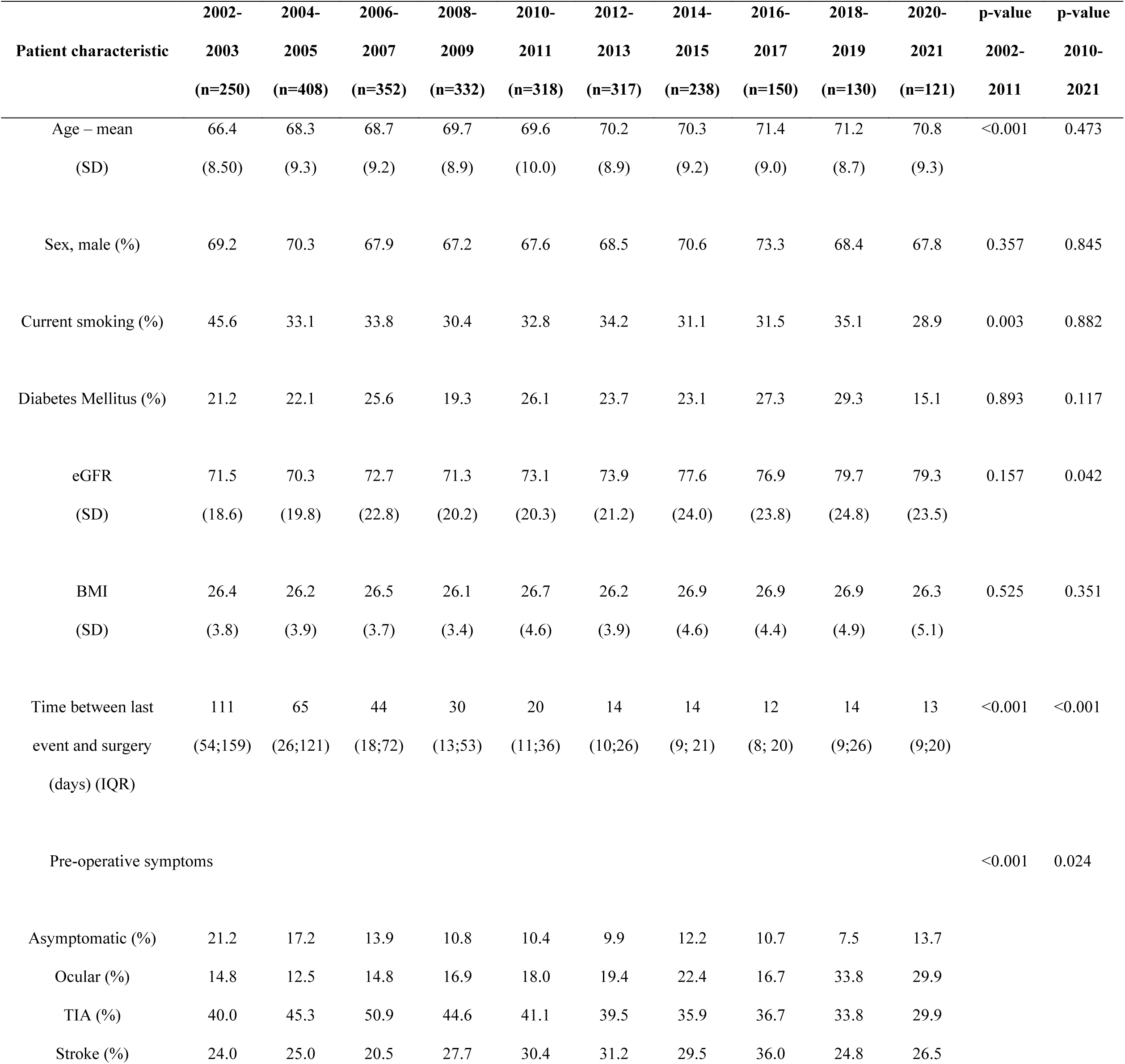
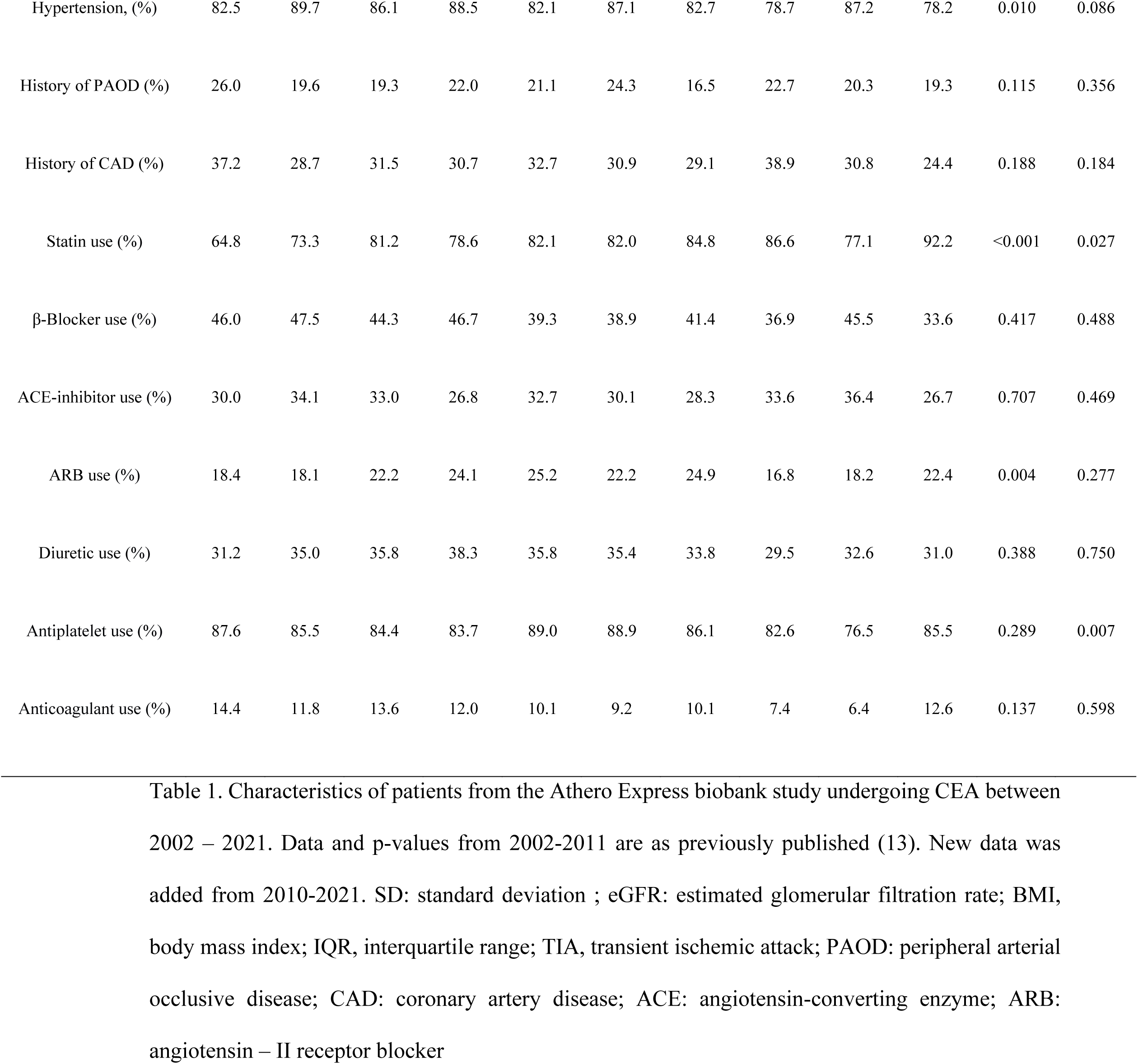
Patient Characteristics Over Time.

Characteristics of patients from the Athero Express biobank study undergoing CEA between 2002 – 2021.
|  | 2002- | 2004- | 2006- | 2008- | 2010- | 2012- | 2014- | 2016- | 2018- | 2020- | p-value | p-value |
| --- | --- | --- | --- | --- | --- | --- | --- | --- | --- | --- | --- | --- |
| Patient characteristic | 2003 | 2005 | 2007 | 2009 | 2011 | 2013 | 2015 | 2017 | 2019 | 2021 | 2002- | 2010- |
|  | (n=250) | (n=408) | (n=352) | (n=332) | (n=318) | (n=317) | (n=238) | (n=150) | (n=130) | (n=121) | 2011 | 2021 |
| Age – mean | 66.4 | 68.3 | 68.7 | 69.7 | 69.6 | 70.2 | 70.3 | 71.4 | 71.2 | 70.8 | <0.001 | 0.473 |
| (SD) | (8.50) | (9.3) | (9.2) | (8.9) | (10.0) | (8.9) | (9.2) | (9.0) | (8.7) | (9.3) |  |  |
| Sex, male (%) | 69.2 | 70.3 | 67.9 | 67.2 | 67.6 | 68.5 | 70.6 | 73.3 | 68.4 | 67.8 | 0.357 | 0.845 |
| Current smoking (%) | 45.6 | 33.1 | 33.8 | 30.4 | 32.8 | 34.2 | 31.1 | 31.5 | 35.1 | 28.9 | 0.003 | 0.882 |
| Diabetes Mellitus (%) | 21.2 | 22.1 | 25.6 | 19.3 | 26.1 | 23.7 | 23.1 | 27.3 | 29.3 | 15.1 | 0.893 | 0.117 |
| eGFR | 71.5 | 70.3 | 72.7 | 71.3 | 73.1 | 73.9 | 77.6 | 76.9 | 79.7 | 79.3 | 0.157 | 0.042 |
| (SD) | (18.6) | (19.8) | (22.8) | (20.2) | (20.3) | (21.2) | (24.0) | (23.8) | (24.8) | (23.5) |  |  |
| BMI | 26.4 | 26.2 | 26.5 | 26.1 | 26.7 | 26.2 | 26.9 | 26.9 | 26.9 | 26.3 | 0.525 | 0.351 |
| (SD) | (3.8) | (3.9) | (3.7) | (3.4) | (4.6) | (3.9) | (4.6) | (4.4) | (4.9) | (5.1) |  |  |
| Time between last event and surgery (days) (IQR) | 111 (54;159) | 65 (26;121) | 44 (18;72) | 30 (13;53) | 20 (11;36) | 14 (10;26) | 14 (9; 21) | 12 (8; 20) | 14 (9;26) | 13 (9;20) | <0.001 | <0.001 |
| Pre-operative symptoms |  |  |  |  |  |  |  |  |  |  | <0.001 | 0.024 |
| Asymptomatic (%) | 21.2 | 17.2 | 13.9 | 10.8 | 10.4 | 9.9 | 12.2 | 10.7 | 7.5 | 13.7 |  |  |
| Ocular (%) | 14.8 | 12.5 | 14.8 | 16.9 | 18.0 | 19.4 | 22.4 | 16.7 | 33.8 | 29.9 |  |  |
| TIA (%) | 40.0 | 45.3 | 50.9 | 44.6 | 41.1 | 39.5 | 35.9 | 36.7 | 33.8 | 29.9 |  |  |
| Stroke (%) | 24.0 | 25.0 | 20.5 | 27.7 | 30.4 | 31.2 | 29.5 | 36.0 | 24.8 | 26.5 |  |  |
| Hypertension, (%) | 82.5 | 89.7 | 86.1 | 88.5 | 82.1 | 87.1 | 82.7 | 78.7 | 87.2 | 78.2 | 0.010 | 0.086 |
| History of PAOD (%) | 26.0 | 19.6 | 19.3 | 22.0 | 21.1 | 24.3 | 16.5 | 22.7 | 20.3 | 19.3 | 0.115 | 0.356 |
| History of CAD (%) | 37.2 | 28.7 | 31.5 | 30.7 | 32.7 | 30.9 | 29.1 | 38.9 | 30.8 | 24.4 | 0.188 | 0.184 |
| Statin use (%) | 64.8 | 73.3 | 81.2 | 78.6 | 82.1 | 82.0 | 84.8 | 86.6 | 77.1 | 92.2 | <0.001 | 0.027 |
| β-Blocker use (%) | 46.0 | 47.5 | 44.3 | 46.7 | 39.3 | 38.9 | 41.4 | 36.9 | 45.5 | 33.6 | 0.417 | 0.488 |
| ACE-inhibitor use (%) | 30.0 | 34.1 | 33.0 | 26.8 | 32.7 | 30.1 | 28.3 | 33.6 | 36.4 | 26.7 | 0.707 | 0.469 |
| ARB use (%) | 18.4 | 18.1 | 22.2 | 24.1 | 25.2 | 22.2 | 24.9 | 16.8 | 18.2 | 22.4 | 0.004 | 0.277 |
| Diuretic use (%) | 31.2 | 35.0 | 35.8 | 38.3 | 35.8 | 35.4 | 33.8 | 29.5 | 32.6 | 31.0 | 0.388 | 0.750 |
| Antiplatelet use (%) | 87.6 | 85.5 | 84.4 | 83.7 | 89.0 | 88.9 | 86.1 | 82.6 | 76.5 | 85.5 | 0.289 | 0.007 |
| Anticoagulant use (%) | 14.4 | 11.8 | 13.6 | 12.0 | 10.1 | 9.2 | 10.1 | 7.4 | 6.4 | 12.6 | 0.137 | 0.598 |

**Table 2.** Presence of Atherosclerotic Plaque Components Over Time.

Presence of different atherosclerotic plaque components of patients undergoing CEA between 2002 – 2021.
| **Atherosclerotic plaque components** | **2002-2003**<br>**(n=250)** | **2004-2005**<br>**(n=408)** | **2006-2007**<br>**(n=352)** | **2008-2009**<br>**(n=332)** | **2010-2011**<br>**(n=318)** | **2012-2013**<br>**(n=317)** | **2014-2015**<br>**(n=238)** | **2016-2017**<br>**(n=150)** | **2018-2019**<br>**(n=133)** | **2020-2021**<br>**(n=121)** | **P value**<br>**2002-2010** | **p-value**<br>**2010-2021** |
| --- | --- | --- | --- | --- | --- | --- | --- | --- | --- | --- | --- | --- |
| Lipid core >40% of plaque, % | 33.2 | 36.0 | 26.7 | 21.1 | 15.8 | 21.7 | 50.0 | 32.8 | 35.2 | 26.2 | <0.001 | <0.001 |
| Intraplaque Hemorrhage, % | 74.4 | 75.5 | 62.5 | 49.1 | 41.4 | 53.8 | 62.6 | 67.7 | 73.6 | 68.9 | <0.001 | <0.001 |
| Collagen, % | 73.6 | 85.8 | 80.1 | 76.8 | 79.0 | 79.4 | 67.4 | 66.7 | 81.2 | 70.5 | 0.643 | 0.111 |
| Macrophages % | 50.0 | 61.5 | 70.8 | 48.3 | 42.6 | 33.5 | 74.3 | 67.2 | 67.2 | 59.6 | <0.001 | <0.001 |
| SMC % | 64.8 | 74.4 | 71.1 | 72.8 | 69.0 | 52.0 | 48.6 | 57.9 | 62.3 | 77.0 | <0.001 | <0.001 |
| Calcifications, % | 51.6 | 62.0 | 60.5 | 51.0 | 24.9 | 23.4 | 26.7 | 39.1 | 46.7 | 36.1 | <0.001 | <0.001 |

### Patients characteristics

Patients’ characteristics in the total period of 2002-2021 undergoing CEA are shown in Table 1. Data and p-values from 2002-2011 were as previously published (13) and data and p-values from 2010-2021 were added. Age at the time of surgery increased between 2002 through 2011 (p<0.001) while age did not differ between 2010 through 2021. Sex was not significantly different over time in both periods while percentage of active smoking patients decreased between 2002-2011 but did not change between 2010-2021. Percentage of diabetes and BMI did not change over time in both periods. Mean eGFR was increasing in 2010-21 from 73.1 in 2010-11 to 79.3 in 2020-21. Mean eGFR was higher in 2010-21 compared with 2002-2011 (78.8 vs 72.0, p<0.001). Median time between last event and surgery was decreasing in both periods: 111 days in 2002 through 2003, 20 days in 2010 through 2011 and 13 days in 2020 through 2021. Median time to event was shorter in 2010-21 compared to 2002-11 (15 days vs 39 days in 2002-11, p<0.001). The mean percentage of asymptomatic patients decreased from 2002 – 2011 to 2010-2021 (14.5% vs 10.7%, p=0.002) while the mean percentage of stroke patients was higher in 2010-21 compared to 2002-2011 (30.1% vs 26.0%, p=0.013). Peripheral arterial occlusive disease (PAOD), hypertension and history of Coronary Artery Disease (CAD) did not change over time in both periods.

### Patients preoperative medication prescription over time

Preoperative medication prescription from 2002-2021 are presented in Table 1. Statin use increased in both periods: 64,8% in 2002 through 2003, 82,1% in 2010 through 2011, 92,2% in 2020 through 2021 with mean statin use being higher in the 2010-21 cohort (83.3% vs 77.0%, <0.001). The use of beta-blockers and angiotensin-converting enzyme inhibitors did not differ over time in both periods. Angiotensin II antagonist usage increased in period 2002-2011 but did not change between 2010-2021. Diuretics and anticoagulant use did not change over time for both periods. Mean anti-platelet usage was slightly higher in the 2002-2011 cohort when compared to the 2010-2021 cohort (88.9% vs 86.0%, p=0.021).

### Patients carotid plaque composition

Plaque histology was performed on the collected atherosclerotic carotid plaques for the presence of the following plaque characteristics: lipid core >40%, intraplaque hemorrhage, collagen, macrophages, SMC’s and calcifications. Results were dichotomized as aforementioned. The percentage of patients having these plaque characteristic are summarized in Table 2. The percentage of patients with a lipid core >40% significantly decreased in the period 2002-2011. In contrast, lipid core >40% increased during the period 2010-2021. A similar pattern was also seen for other destabilizing plaque characteristics: intraplaque hemorrhage, macrophages and calcification with a decrease over time in the period 2002-2011 but now an increase over time in the period 2010-2021. On the other hand, for the stabilizing plaque characteristics, plaque collagen did not change over time in the period 2002-2011 but showed a tendency to decline over time for the period 2010-2021. SMC presence increased over time for both the period 2002-2011 and 2010-2021.

### Change in semiquantitative plaque characteristics between 2010-2021

Having found changes over time in the period 2010-2021 in the dichotomous results for plaque characteristics, we investigated how semiquantitative information based on the degree of staining changed in this period (Figure 1). The number of plaques containing >40% fat is increased (upper left). Plaques with no staining of thrombus is low in most recent years (upper right). The number of plaques with no or minor collagen and no or minor SMC’s staining showed little change in the last years (left middle and lower) when compared to the older time intervals. Moderate and heavy macrophage staining increased over time (middle right). For calcification, moderate and heavy staining was more often present in recent time intervals (lower right).

**Figure 1.**
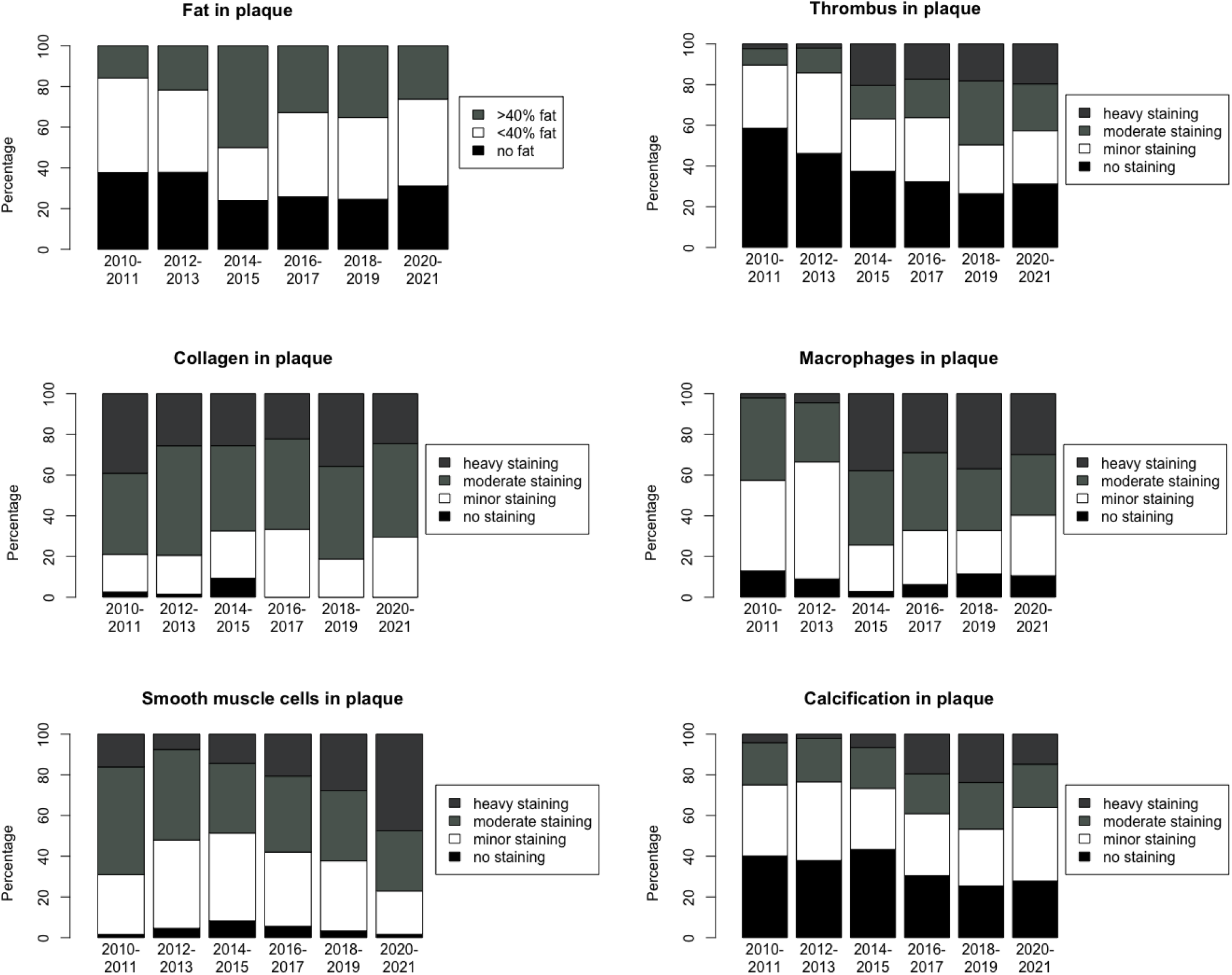
Semiquantitative plaque information from 2010 and 2021 based on the degree of staining.

### Adjusted odds ratios for the presence of atherosclerotic plaque characteristics, per 2-year increase from 2010 to 2021

To explore if changes over time remained after correction for confounders, we assessed the association between plaque characteristics and year of surgery (Figure 2) after adjustment for patient characteristics that changed over time (diabetes mellitus, eGFR, time between last event and surgery, symptoms, hypertension, history of CAD, statins use and antiplatelet drug use). After adjustment, all of the aforementioned plaque characteristics, except for SMC and collagen content, remained significantly associated with year of surgery and indicated an increase in features that are considered to be associated with carotid plaque instability over time. Adjusted odds ratio’s per 2-year increase: calcification 1,204 (95% confidence interval, 1,090-1,331; p<0,001), intraplaque hemorrhage 1,358 (95% confidence interval, 1,230-1,502; p<0,001); >40% fat in the plaque 1,271 (95% confidence interval, 1,150-1,407; p<0,001); macrophages 1,366 (95% confidence interval, 1,238-1,511; p<0,001). See figure 3 for histological sections of the aforementioned plaque characteristics.

**Figure 2.**
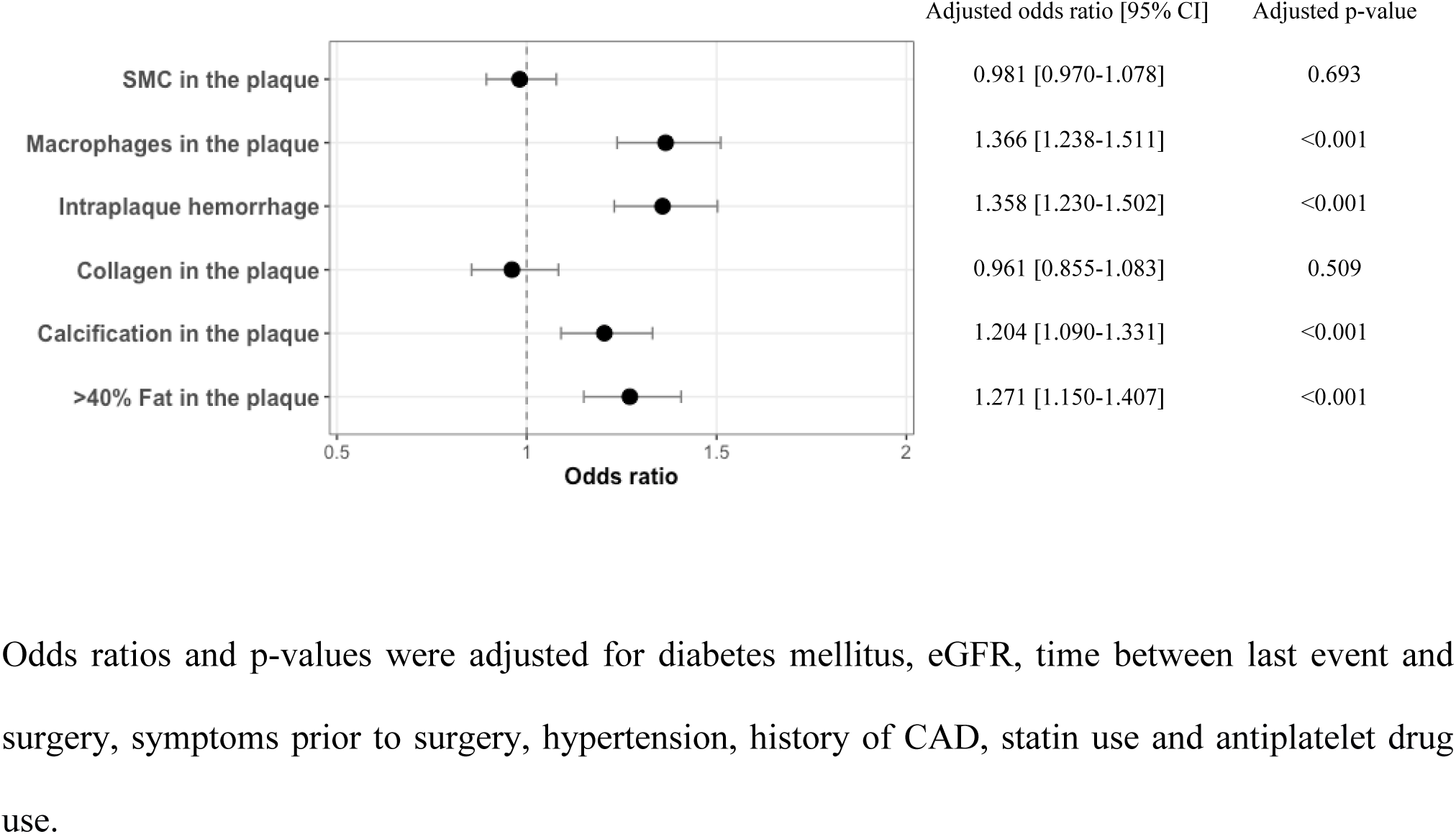
Adjusted odds ratios for the presence of atherosclerotic plaque characteristics, per 2-year increase in time from 2010 to 2021.

**Figure 3.**
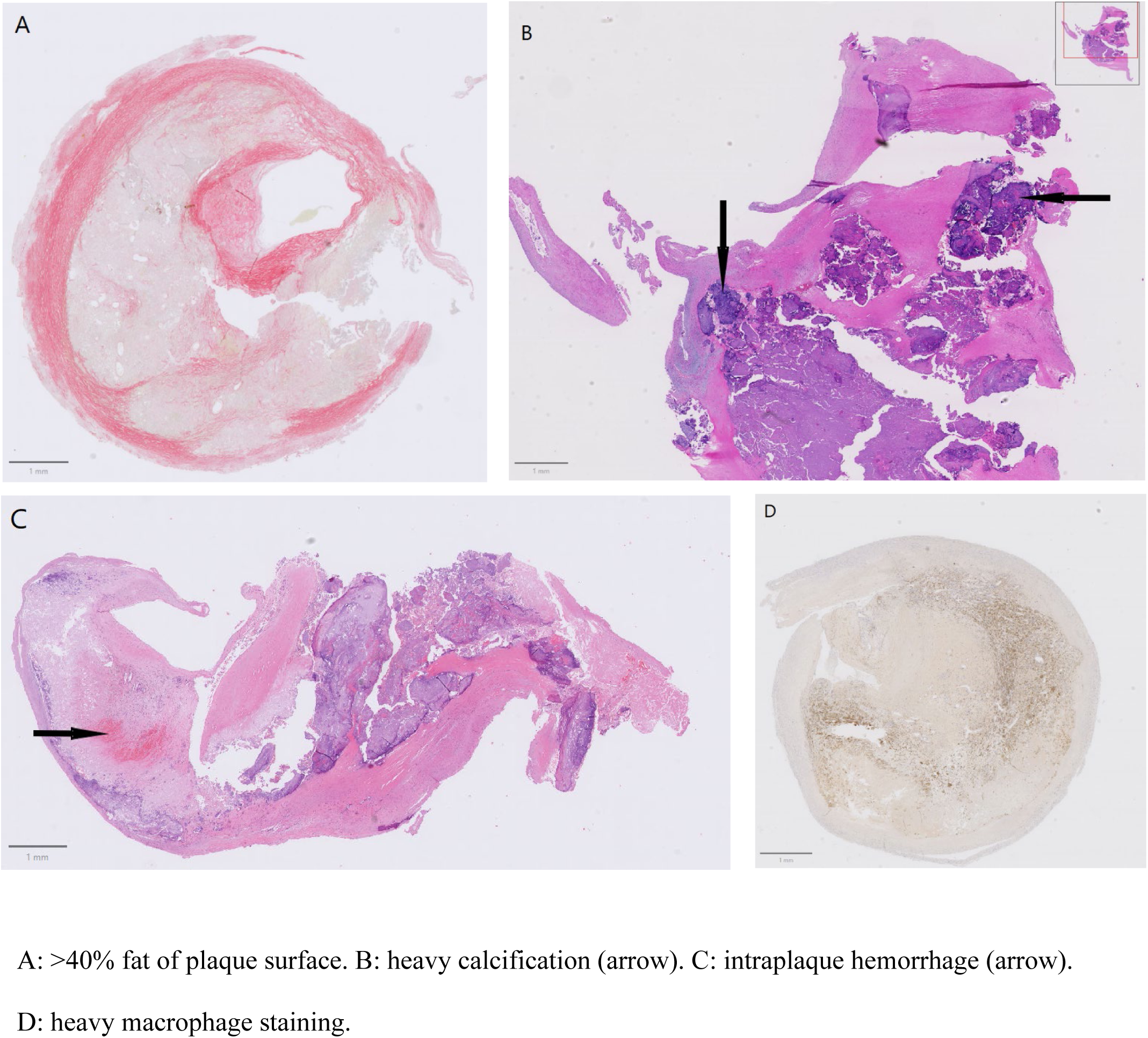
Histological sections of adverse plaque characteristics.

### Carotid plaque composition per preoperative symptom subgroup prior to CEA

Since plaques from asymptomatic patients have a more fibrous and less inflammatory phenotype as compared with patients with transient ischemic attack and stroke (21), we performed a sub-analysis on change of plaque characteristics over time in patients with different symptoms (Table 3). This table shows for each patient subgroup from 2010 to 2021 the same significant increase of adverse plaque characteristics: presence of a lipid core >40%, intraplaque hemorrhage and macrophages. Calcifications changed significantly over time in the asymptomatic and ocular group but did not change in the TIA and stroke patient subgroup.

**Table 3.**
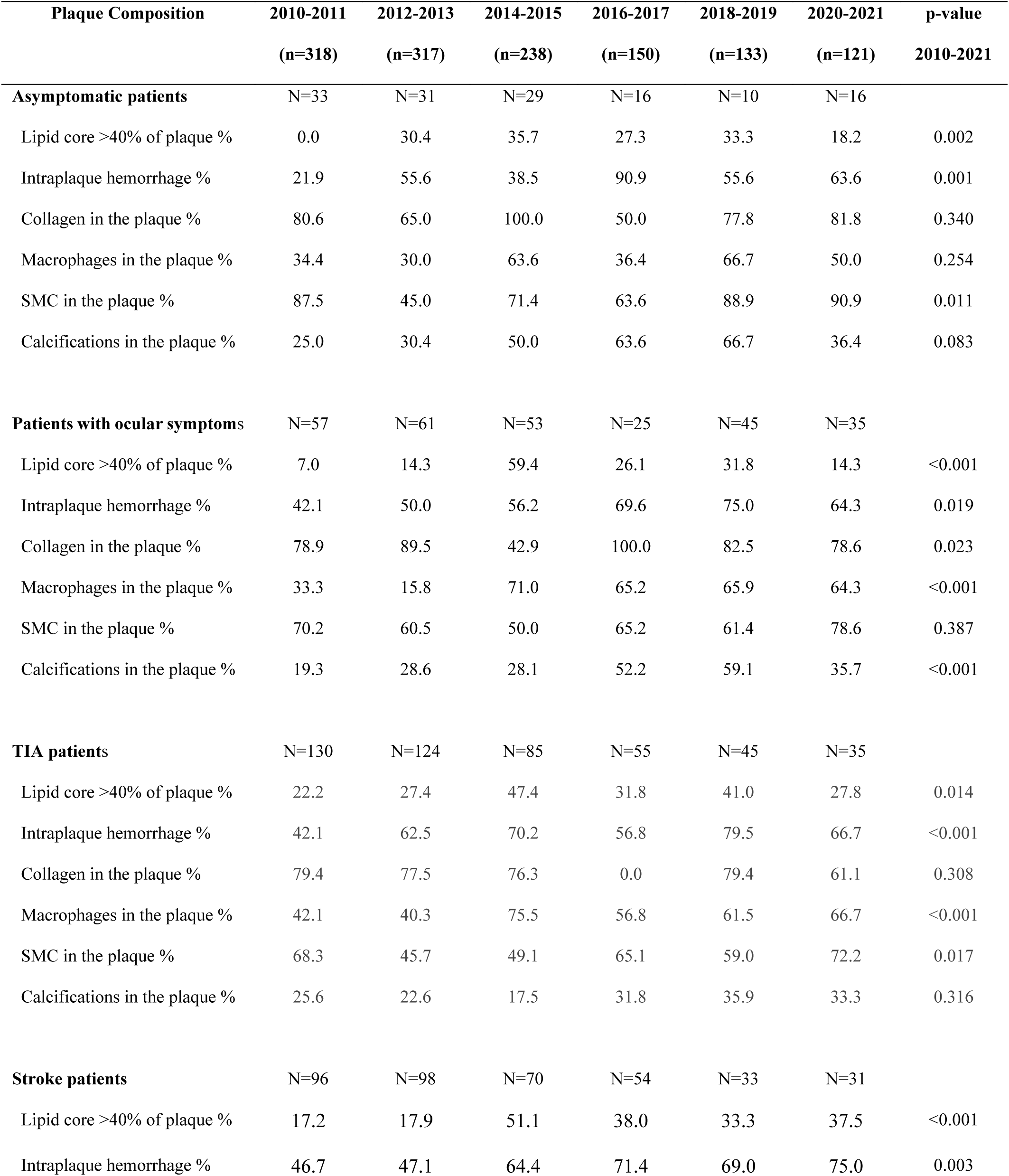
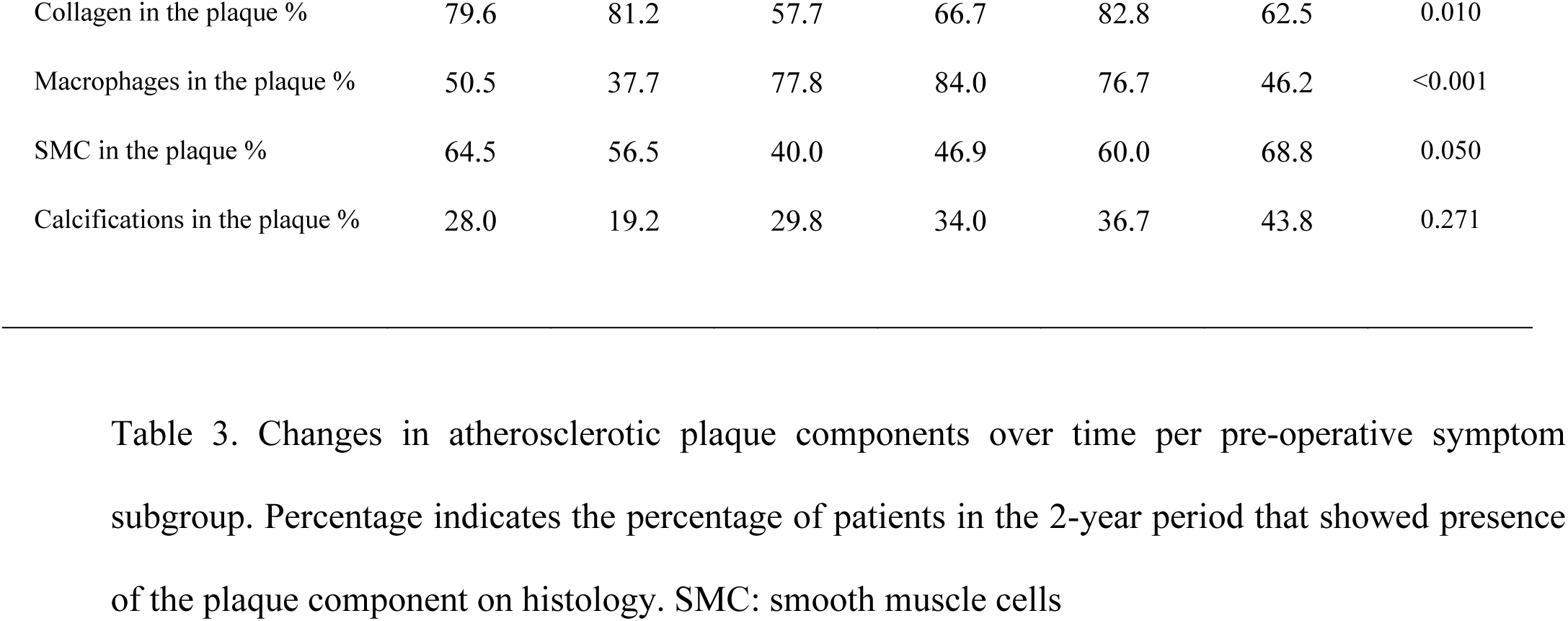
Carotid Plaque Composition per pre-operative symptom subgroup.

## Discussion

Although world-wide mortality of ASCVD is increasing, in Western-Europe prevalence remained unchanged, ASCVD mortality declined until 2013 and hospital admission and ASCVD prescriptions increased.(11) Similarly, vulnerable plaque characteristics in human carotid plaques were decreasing between 2002-2011.(13) Time dependent changes in plaque characteristics in the last decade, however, are unknown. We have now shown that atherosclerotic plaques of carotid endarterectomy patients are not further stabilising as was shown until 2011. In contrast, vulnerable plaque characteristics such as macrophages, calcification, intraplaque hemorrhage and fat are increasing over time since 2010.

### Patient characteristics over time

In the last decade, the age of CEA patients in our study did not increase while also the decline in smoking stabilized. The prevalence of diabetes showed a tendency towards an increase, as did patients in the SWEDEHEART registry (12) and in the male elderly Dutch population between 2010-2020 (22) which forms the majority of our patients studied. In the COVID19 year 2020-21, however, the percentage of diabetes patients is the lowest since 2002. Despite a tendency of an increase in diabetes, eGFR is increasing, suggesting that kidney function of these CEA patients is improving.

A major change is seen in the decrease in time between event and operation. This is expected as guidelines recommend that surgeons should strive to operate within 14 days after the index neurologic event. (23,24) It is, however, known that early after stroke and TIA, carotid plaques have a more unstable plaque phenotype but stabilize in the weeks after the event.(25)

Medication use showed that pre-operative statin use increased in both the 2002-2011 and 2010-2021 period. The increase in Angiotensin II antagonists in 2002-2011 is stabilized in the 2010-2021 period while antiplatelet was slightly lower in 2010-21. Other pre-operative medication use has not changed significantly. Although targeting the same pathways and pathologies, we have to realize that in the last decade other specific medication prescriptions within the same drug entity and/or combined treatments of more than 1 pathway might have been used, resulting in a change in the percentage of patients treated.

### Plaque characteristics over time

This study shows time-dependent changes towards plaques with characteristics thought to be associated with plaque vulnerability. This seems to be in contrast with the decrease in mortality in Western Europe. (10) Around 23,000 strokes occur each year in the Netherlands and estimates indicate that the prevalence of stroke has risen over the past ten years(26). In our study, we also saw an increase of symptomatic patients and a decrease in asymptomatic patients in the last decade. TIA and stroke before CEA have been associated with a more vulnerable plaque phenotype compared to asymptomatic patients (7,8). The decrease in time to surgery and increase in symptomatic patients could be an explanation for the increase in plaque characteristics associated with vulnerability, however, the increase in vulnerable plaque characteristics remained significant after correction of these confounders. Next to this, subgroup analysis of plaque characteristics in the four different pre-operative symptom groups shows the same increase in vulnerable plaque characteristics in each of these groups.

### Vulnerable plaque characteristics and cardiovascular events

The vulnerable plaque with a large atheroma, heavy macrophage deposition, covered by a thin fibrous cap was considered to be prone for rupture and defined as the thin-capped fibroatheroma (TCFA). (27) This concept is currently challenged (28), mainly since only a minority of TCFA plaques lead to events. (2). Also in this study, we did not find any association between vulnerable plaque characteristics and cardiovascular events over time (suppl data S1). Of the vulnerable plaque characteristics of fat, calcification, macrophages and intraplaque hemorrhage that significantly increased in time, only intraplaque hemorrhage has been previously associated with future cardiovascular events (1). Previously, the time associated decrease between 2002-2011 in plaque characteristics believed to be associated with plaque vulnerability including intraplaque hemorrhage were also not associated with a reduced event rate in this cohort of CEA patients. (13) This is in line with previous studies suggesting a limited role of plaque characteristics in the occurrence of future cardiovascular events (1,2,28).

In the 2002-2011 study (13), the decrease in these vulnerable plaque characteristics coincided with an increase in statin use, suggesting statin use might be an explanation of a more stable plaque characteristics. Besides their primary lipid lowering effect, statins exert anti-inflammatory effects in cardiovascular patients. (29) The pleiotropic immune-modulatory properties of statins have gained much attention in the last decade as additional explanation of reduction in cardiovascular events. (30) In contrast, we now find more vulnerable plaque characteristics but find also a higher statin use in the 2010-2021 cohort as compared to the 2002-2010 cohort. Several reasons might be underlying this apparent contradiction. Firstly, we showed in the Athero Express cohort that statin use was associated with more histological presence of plaque macrophages (31). But statin use is, in turn, also associated with less metabolic activity of plaque macrophages. (32) Secondly, intraplaque hemorrhage and plaque vessel density in men are the only plaque characteristics associated with recurrent cardiovascular events, but the majority of plaque characteristics (e.g. macrophages, atheroma, collagen and SMC’s) are not.(1) Thirdly, in this study, we describe more vulnerable plaque characteristics compared to the previous decade while there is no association with recurrent events (suppl data Fig 1) and a decrease in cardiovascular mortality in western Europe in the background. (10) This suggests that histological quantification of plaque characteristics are of limited value to predict atherosclerotic progression and future events and should encompass metabolic activity as well.

### Limitations

This CEA cohort might not represent other ASCVD patients. Our observations may have been influenced by other cardiovascular risk factors as we do not have data on physical activity or additional drug use as only data on medication presented are available. Regarding limitations, bias in studying time-dependent trends might arise if, for instance, the disease diagnosis criteria have changed. In the Athero Express study, we did not notice a reduction in the total number of patients. Nonetheless, we cannot rule out that the variations in our observations over time may be due to changes in patient characteristics and surgical indications that occurred during the same period.

## Conclusion

In conclusion, we found that in the last decade carotid plaque characteristics did not show the same trend of stabilisation as the cohort from 2002 to 2010. Moreover, plaque characteristics associated with plaque vulnerability are increased in this large cohort of CEA patients. With a background of decreasing ASCVD mortality in Western-Europe and no relation with cardiovascular events, these time dependent changes are evident but not completely understood or explained by traditional histological measures of atherosclerotic risk. This asks for a continuing search of other factors that not just associate with the risk for an event but also with specific plaque types underlying these events.

## Data Availability

All data produced in the present work are contained in the manuscript

## Acknowledgments

None

## Supplemental Material

Figure S1

## Non-standard Abbreviations and Acronyms

ASCVD: atherosclerotic cardiovascular disease
CAD: coronary artery disease
CEA: Carotid endarterectomy
HE: haematoxylin-eosin
MDRD: Modification of Diet in Renal Disease formula
PAOD: peripheral arterial occlusive disease
SMC: smooth muscle cell
TCFA: thin-capped fibroatheroma
TIA: transient ischemic attack

## Supplemental data

**Figure S1.**
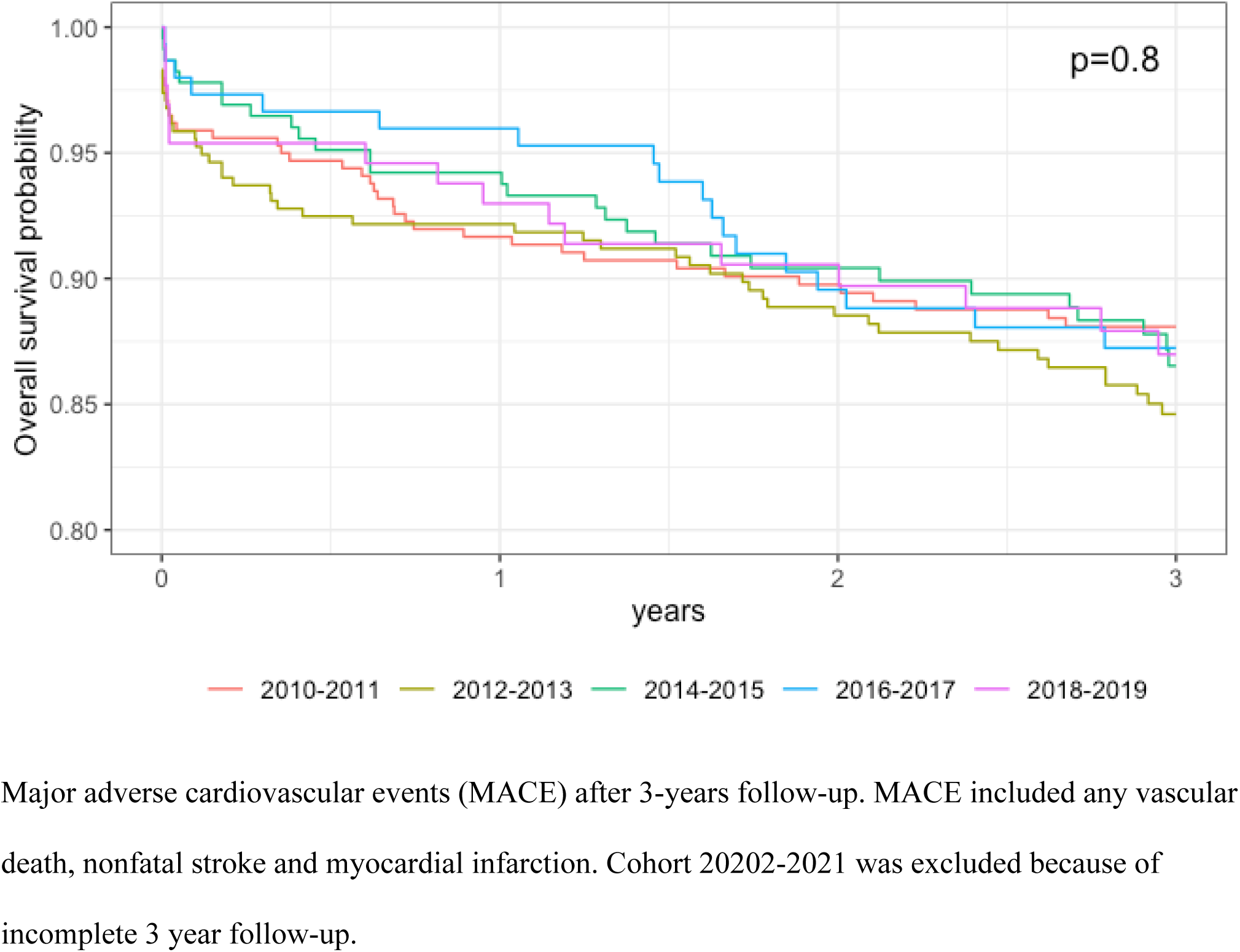
Kaplan Meier survival curves of the patients included in the Athero-Express, stratified by inclusion year (2010-2021).

